# Interscalene Block Enhances Wound Healing, Modulates Immune Response, and Accelerates Recovery After Shoulder Surgery

**DOI:** 10.1101/2025.11.13.25340182

**Authors:** Arzu Esen Tekeli, Cihan Adanaş, Nureddin Yüzkat, Mehmet Parlak, Sanjib Das Adhikary, Harun Kiyat, Mine Uzlaş

**Affiliations:** Research Scholar, Department of Anesthesiology and Perioperative Medicine, University of Virginia, Virginia, USA; Department of Orthopedics and Traumatology, Van Yuzuncu Yil University, School of Medicine, Van, TURKEY; Department of Anesthesiology and Reanimation, Van Yuzuncu Yil University, School of Medicine, Van, TURKEY; Department of Medical Microbiology, Van Yuzuncu Yil University School of Medicine, Van, TURKEY; Department of Anesthesiology and Perioperative Medicine, Penn State College of Medicine, Pennsylvania, USA

**Author notes:** Corresponding Author: E mail.

**Keywords:** Interscalene block, shoulder surgery, wound healing, immune modulation, postoperative recovery, regional anesthesia

## Abstract

**Background:** Shoulder surgeries are among the most painful orthopedic procedures, and optimal perioperative management is crucial for recovery. Interscalene block (ISB) is widely used for analgesia, yet its effects on wound healing and postoperative immune function remain poorly defined. This study aimed to determine whether ISB enhances wound healing, modulates immune responses, and improves recovery outcomes compared with general anesthesia (GA).

**Methods:** This study is a prospective, randomized, controlled clinical trial designed to evaluate the effects of interscalene block on wound healing and postoperative immune response in patients undergoing elective open shoulder surgery.Thirty-four patients undergoing elective rotator cuff repair were randomly assigned to GA (n = 17) or ISB (n = 17) via sealed-envelope randomization. Wound healing was assessed using the Toronto Symptom Assessment System for Wounds (TSAS-W) on postoperative days 5 and 14. Immune response was evaluated by platelet count and serum levels of IL-1, IL-2, TNF-α, EGF, TGF-β, and PDGF at baseline, 24, and 48 hours postoperatively. Secondary outcomes included postoperative pain (VAS), mobilization times, analgesic consumption, and incidence of nausea and vomiting. Statistical analyses incorporated parametric and non-parametric tests, repeated measures analysis, chi-square/Fisher’s exact tests, Bonferroni correction, and effect size calculations (p < 0.05).

**Results:** ISB significantly accelerated wound healing, with lower TSAS-W scores on days 5 and 14 compared with GA (p < 0.05). Postoperative platelet counts and levels of IL-1, IL-2, EGF, TGF-β, and PDGF were significantly higher in the ISB group at 24 and 48 hours (p < 0.05), whereas TNF-α remained unchanged. ISB also provided superior analgesia, reflected by lower VAS scores, reduced analgesic consumption, faster mobilization, and decreased incidence of postoperative nausea and vomiting (all p < 0.05).

**Conclusion:** Interscalene block not only provides effective analgesia but also enhances wound healing and modulates immune function, promoting faster and safer recovery after shoulder surgery. These findings support ISB as a multifaceted perioperative intervention with benefits beyond pain control.

## Introduction

Interscalene block is a safe and effective method of anesthesia and analgesia for patients undergoing shoulder procedures such as rotator cuff repair, decompression, and arthroplasty (1). The selection of an appropriate anesthetic technique is equally as important as the surgical procedure itself in ensuring patient comfort and minimizing postoperative pain (2). Shoulder surgeries often require active patient participation to assess joint mobility, stability, and function. Therefore, an anesthesia approach that facilitates both patient cooperation during the procedure and optimal comfort is of critical importance (3,4). Shoulder surgeries are among the most painful interventions in orthopedic practice. Concerns regarding pain and elevated postoperative pain levels can lead to delayed mobilization and permanent limitations in range of motion. Therefore, brachial plexus blockade has been widely recognized as an effective method of providing analgesia (5). The shoulder joint is primarily innervated by the suprascapular and axillary nerves, which originate from the brachial plexus. Interscalene brachial plexus block typically targets the C5–C7 nerve roots (6). Historically, peripheral nerve blocks were performed using surface anatomical landmarks and nerve stimulation techniques. Today, ultrasound (US) guidance has become the standard in the practice of peripheral nerve blocks. As it allows real-time visualization of the needle and surrounding anatomy, leading to increased efficacy and block success rates, shorter performance and onset times, improved postoperative pain control, faster recovery, and a reduced risk of complications (7).

In nerve block procedures, local anesthetic agents are administered either alone or in combination with adjuvants. These agents function as voltage-gated sodium channel blockers applied to nerves for anesthetic or analgesic purposes. In vitro studies have demonstrated that local anesthetics exert direct effects on wound healing and cancer cells through multiple mechanisms, including the regulation of cell proliferation, apoptosis, migration, and invasion (8,9). Wound healing itself is an evolutionarily conserved, complex, multicellular process aimed at restoring the barrier function of the skin. This process involves the coordinated efforts of various cell types, including keratinocytes, fibroblasts, endothelial cells, macrophages, and platelets. The migration, infiltration, proliferation, and differentiation of these cells culminate in an inflammatory response, the formation of new tissue, and ultimately, wound closure. This complex sequence of events is orchestrated and regulated by an equally intricate signaling network comprising numerous growth factors, cytokines, and chemokines (10). Interleukin-1 (IL-1), Interleukin-2 (IL-2), tumor necrosis factor-alpha (TNF-α), epidermal growth factor (EGF), transforming growth factor-beta (TGF-β), and platelet-derived growth factor (PDGF) are among the key cytokines with significant roles in the wound healing process.

Interleukin-1 (IL-1) is a proinflammatory cytokine that influences cellular and organ-level inflammatory responses, immune reactions, and the regulation of homeostasis (11). Interleukin-2 (IL-2), the first cytokine to be molecularly cloned, has been shown to be an essential T cell growth factor required for the proliferation of T cells and the generation of effector and memory cells (12). Tumor necrosis factor-alpha (TNF-α) is a proinflammatory cytokine that plays a direct role in the pathogenesis of various acute disease conditions, including sepsis, traumatic injury, cardiac and cerebral ischemia, asthma, and burns TNF-α and inflammatory mediators can regulate cell apoptosis. (13). Epidermal growth factor (EGF) is a key signaling molecule involved in tissue repair and the preservation of tissue equilibrium by modulating cellular viability, expansion, motility, and specialization (14). Transforming growth factor-beta (TGF-β) is a pleiotropic cytokine that influences cellular proliferation, differentiation, programmed cell death, migration, extracellular matrix synthesis, blood vessel formation, and immune cell function. Interestingly, TGF-β exhibits a dual role by acting as a tumor suppressor under certain conditions (15). The platelet-derived growth factor (PDGF) family contributes to numerous cellular growth mechanisms and plays a crucial role in modulating angiogenesis and the development of mesenchymal cells. Consequently, PDGF holds promise as a potential therapeutic agent in the treatment of heart failure (16). In recent years, substantial evidence has been obtained regarding the efficacy of these agents not only in promoting wound healing and epithelialization, but also in the treatment of various autoimmune diseases and certain types of cancer (12,13,16). Various scales have been developed to assess wound healing. Currently, the Toronto Symptom Assessment System for Wounds (TSAS-W) is a novel tool designed to systematically evaluate wound-associated pain and polysymptom burden. TSAS-W consists of 10 symptom parameters (e.g., wound location, classification, size, pain, drainage, odor, etc.), each rated individually on an 11-point numerical rating scale (0–10). Lower scores indicate more favorable healing outcomes. TSAS-W is concise, easy to administer and complete, and its design allows for application across all wound types and classifications, making it a versatile assessment tool (17).

Building upon these findings, the present study aims to evaluate the effects of interscalene block—a widely used technique in anesthesia practice—on platelet count, chemokine levels, wound healing, postoperative pain scores, analgesic consumption, and the incidence of postoperative nausea and vomiting. Our goal is to present and discuss these outcomes within the context of the existing literature.

## Method

### Study design and participation

This prospective, randomized, controlled study was conducted as a two-center trial by Van Yüzüncü Yıl University Faculty of Medicine and Penn State College of Medicine. Patient enrollment was carried out by researchers at Van Yüzüncü Yıl University Faculty of Medicine, while statistical analysis and manuscript preparation were performed with contributions from researchers at Penn State College of Medicine. The study was approved by the Clinical Research Ethics Committee of Van Yüzüncü Yıl University (13.06.2022-14). Following ethical approval, the study was registered on ClinicalTrials. gov (NCT05499897). After the completion of registration, patients who were informed about the study and provided both written and verbal consent were enrolled. Patient personal data cannot be shared due to patient confidentiality. Access to such data may be requested from the Department of Anesthesiology, Faculty of Medicine, Van Yüzüncü Yıl University.

### Inclusion criteria

Patients aged between 18 and 65 years, with ASA physical status I–II, scheduled to undergo elective open shoulder surgery for rotator cuff tear, and who consented to participate in the study.

**Exclusion criteria** are pregnancy, bleeding diathesis, chronic renal insufficiency, liver dysfunction, immunosuppression, intraoperative blood transfusion requirement, emergency or bleeding, severe cardiopulmonary disease, neuropsychiatric disorders, allergy to local anesthetics, morbidly obesity(BMI ≥ 35 kg/m²).

### Sample size

Participants were randomly assigned to either the general anesthesia group or the interscalene block group using the sealed-envelope method (Figure 1). An a priori power analysis for two independent groups (n = 17 per group; total n = 34) was done. Under a two-tailed independent samples t-test with α = 0.05 and power = 0.80, the minimum detectable effect size (Cohen’s d) is approximately 0.96. This implies that the study is adequately powered to detect very large between-group differences, but may be underpowered for detecting small to moderate effects. Consequently, the potential for a Type II error should be considered when interpreting nonsignificant results.

**Figure 1:**
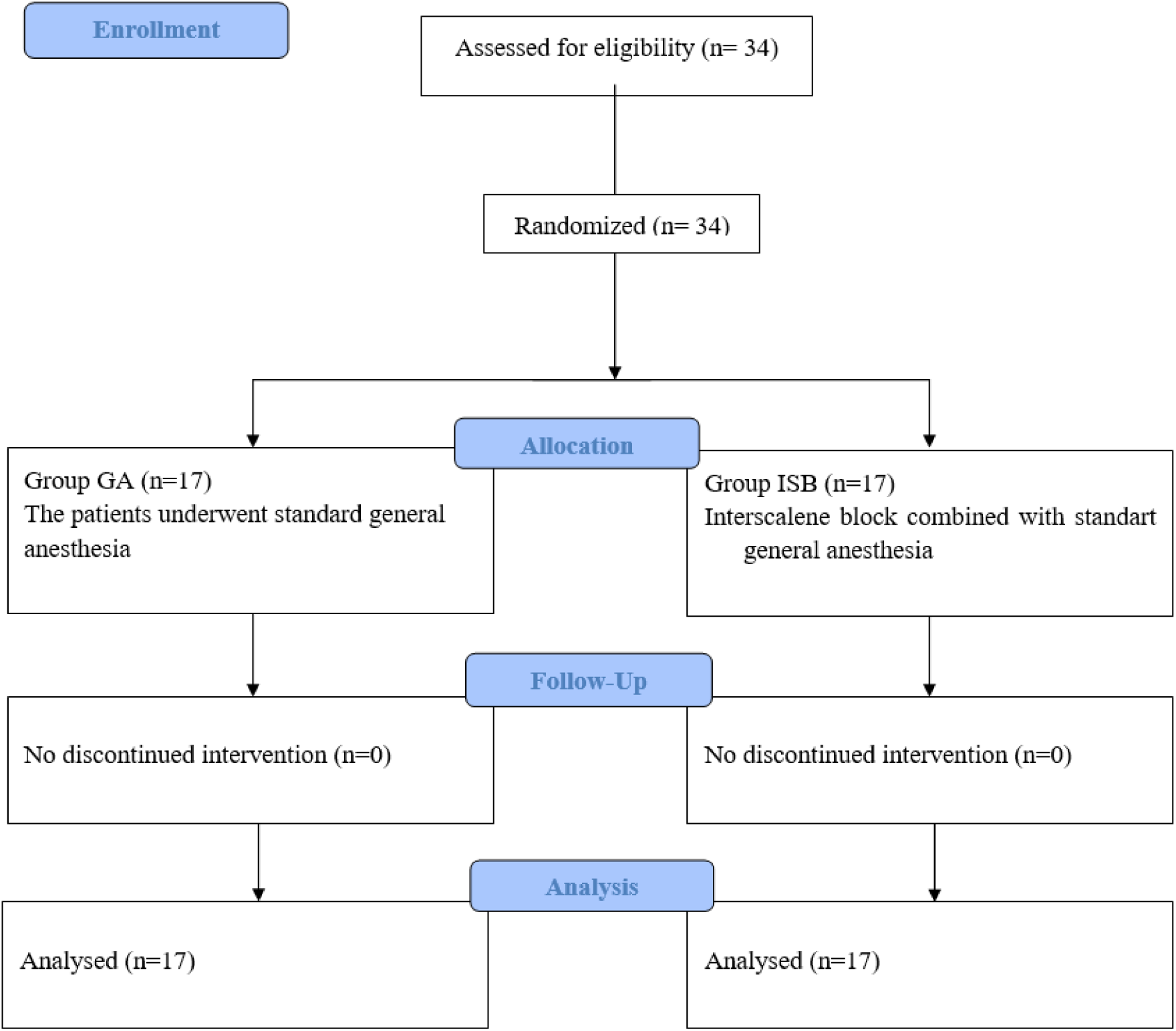
Follow Diagram.

#### Group General Anesthesia (Group GA)

Standard ASA monitoring (non-invasive blood pressure, heart rate, peripheral oxygen saturation, and end-tidal CO₂) was applied. For anesthesia induction 2 mg/kg propofol iv, 0.6 mg/kg rocuronium bromide iv, and 1 µg/kg fentanyl iv was achieved. Anesthesia was maintained with 2 MAC sevoflurane in a mixture of 40% air at a flow rate of 2 L/min. After the surgical procedure, which lasted approximately 50 minutes, 1 mg/kg tramadol iv and 15 mg/kg paracetamol iv were administered 10 minutes prior to extubation for postoperative analgesia in both groups. Postoperative follow-up continued for 48 hours in the Orthopedics and Traumatology ward. For wound healing assessment, patients were recalled for follow-up visits on postoperative days 5 and 14, during which TSAS-W scores were recorded.

#### Group Interscalene Block (Group ISB)

Following the ASA monitoring patients were placed in the supine position with their heads turned toward the unaffected side. Standard skin antisepsis and sterile draping were performed. Ultrasound imaging commenced at the supraclavicular fossa using a 15–6 MHz linear transducer (POCUS Mindray M7 Portable US, Shenzhen, China) to visualize the subclavian artery and brachial plexus. The transducer was then advanced in the cranial direction to identify the C5–6 nerve roots. No sedative agents were administered to patients receiving ISB, and local anesthetic was not applied at the needle insertion site. A 22-gauge 8 cm needle was introduced via a lateral-to-medial approach into the middle scalene muscle and advanced until it was located adjacent to the nerve roots. The needle was consistently visualized using ultrasound to avoid unintentional intraneural or intravascular injection. Upon confirmation of negative blood aspiration, 20 mL of 0.25% bupivacaine was administered around the nerve roots unilaterally. If blood aspiration or radiating pain was encountered during injection, the needle was repositioned under ultrasound guidance. The interscalene block procedure was performed by the same anesthesiologist for all patients. The block was considered successful and adequate for shoulder surgery when anesthesia was achieved in the C4–C6 dermatomes and weakness in elbow flexion was observed. After confirmation, the surgical team was informed that the patient was ready for the procedure.

General anesthesia and interscalene block procedures were performed by two different anesthesiologists, while all surgical interventions were carried out by the same orthopedic surgeon. The study was conducted in a single-blind manner: the anesthesiologist performing the interscalene block was aware of group allocation, whereas both the surgeon and the patient were blinded to the assigned intervention. Blinding was achieved by ensuring that the surgical team did not participate in the block procedure and by keeping patients unaware of which anesthesia technique they received. Both groups underwent identical preoperative preparation, monitoring, surgical procedures, and postoperative care, with the only difference being the method of anesthesia, thereby maintaining procedural similarity between groups.

Platelet count, IL-1, IL-2, TNF-α, EGF, TGF-β, and PDGF levels were assessed preoperatively, at 24 hours, and 48 hours postoperatively, with the results recorded. Patients who were stable after 48 hours were discharged and scheduled for follow-up visits on postoperative days 5 and 14 for wound healing assessment. TSAS-W was used to evaluate wound healing.

The primary outcome of this study was to evaluate the impact of interscalene block on wound healing and postoperative immune response. Wound healing was assessed using the TSAS-W score, while immune response was evaluated through measurement of platelet count and serum concentrations of IL-1, IL-2, TNF-α, EGF, TGF-β, and PDGF. Secondary outcomes were designed to capture clinically relevant aspects of postoperative recovery, including pain intensity, functional recovery, analgesic requirements, and the incidence of nausea and vomiting. Pain intensity was quantified using the Visual Analog Scale (VAS), functional recovery was evaluated by mobilization times, and analgesic consumption and postoperative nausea and vomiting rates were systematically recorded between the two groups.

### Statistical Analysis

Descriptive statistics were presented as mean ± standard deviation (SD) for normally distributed variables, and median (minimum–maximum) for non-normally distributed variables. Categorical variables were expressed as frequencies and percentages. The normality of continuous variables was assessed using the Shapiro–Wilk test. For independent variables, the independent samples t-test was applied for normally distributed data, and the Mann–Whitney U test for non-normal data. Paired continuous variables were analyzed using the paired t-test or Wilcoxon signed-rank test as appropriate. Repeated measures, such as serial VAS scores or immune parameters, were evaluated using repeated measures ANOVA or the Friedman test. Categorical variables were analyzed with the chi-square test or Fisher’s exact test when expected frequencies were low. Bonferroni correction was applied for multiple comparisons where necessary. Effect sizes (Cohen’s d or r) were calculated to assess the magnitude of differences. All statistical analyses were performed using IBM SPSS Statistics for Windows, version 27.0 (IBM Corp., Armonk, NY, USA), and statistical significance was set at p < 0.05.

## Results

No statistically significant differences were found between the groups in terms of demographic data, ASA scores, and smoking status (p > 0.05) (Table 1).

**Table 1:** Demographics.

|  |  | Group GA (n:17) |  |  | Group ISB (n:17) |  |  | p |  |
| --- | --- | --- | --- | --- | --- | --- | --- | --- | --- |
|  |  | Mean±Sd/n-% |  |  | Mean.±Sd/n-% |  |  |  |  |
| **Age** |  | 52.5 | ± | 14.1 | 49.8 | ± | 14.4 | 0.398 | <sup>m</sup> |
| **Height** |  | 165.8 | ± | 8.9 | 164.7 | ± | 5.6 | 0.892 | <sup>m</sup> |
| **Weight** |  | 77.6 | ± | 9.7 | 74.3 | ± | 10.2 | 0.364 | <sup>t</sup> |
| **BMI** |  | 28.2 | ± | 2.9 | 26.9 | ± | 3.3 | 0.227 | <sup>t</sup> |
| **ASA** | **I** | 4 |  | 21.1% | 10 |  | 76.9% | 0.467 | <sup>X²</sup> |
|  | **II** | 13 |  | 68.4% | 7 |  | 53.8% |  |  |
| **Smoking** | **(-)** | 14 |  | 73.7% | 11 |  | 84.6% | 0.975 | <sup>X²</sup> |
|  | **(+)** | 3 |  | 15.8% | 6 |  | 46.2% |  |  |

<sup>t</sup> Independent sample t test / <sup>m</sup> Mann-whitney u test / <sup>X²</sup> Chi-square test

The TSAS-W score was used as the wound healing evaluation score (Figure 2). In the interscalene group, the TSAS-W score on postoperative day 5 and 14 was significantly lower compared to the general anesthesia group (p < 0.05). In both groups, the TSAS-W score on postoperative day 14 was significantly lower than the score on postoperative day 5 (p < 0.05) (Table 2).

**Figure 2:**
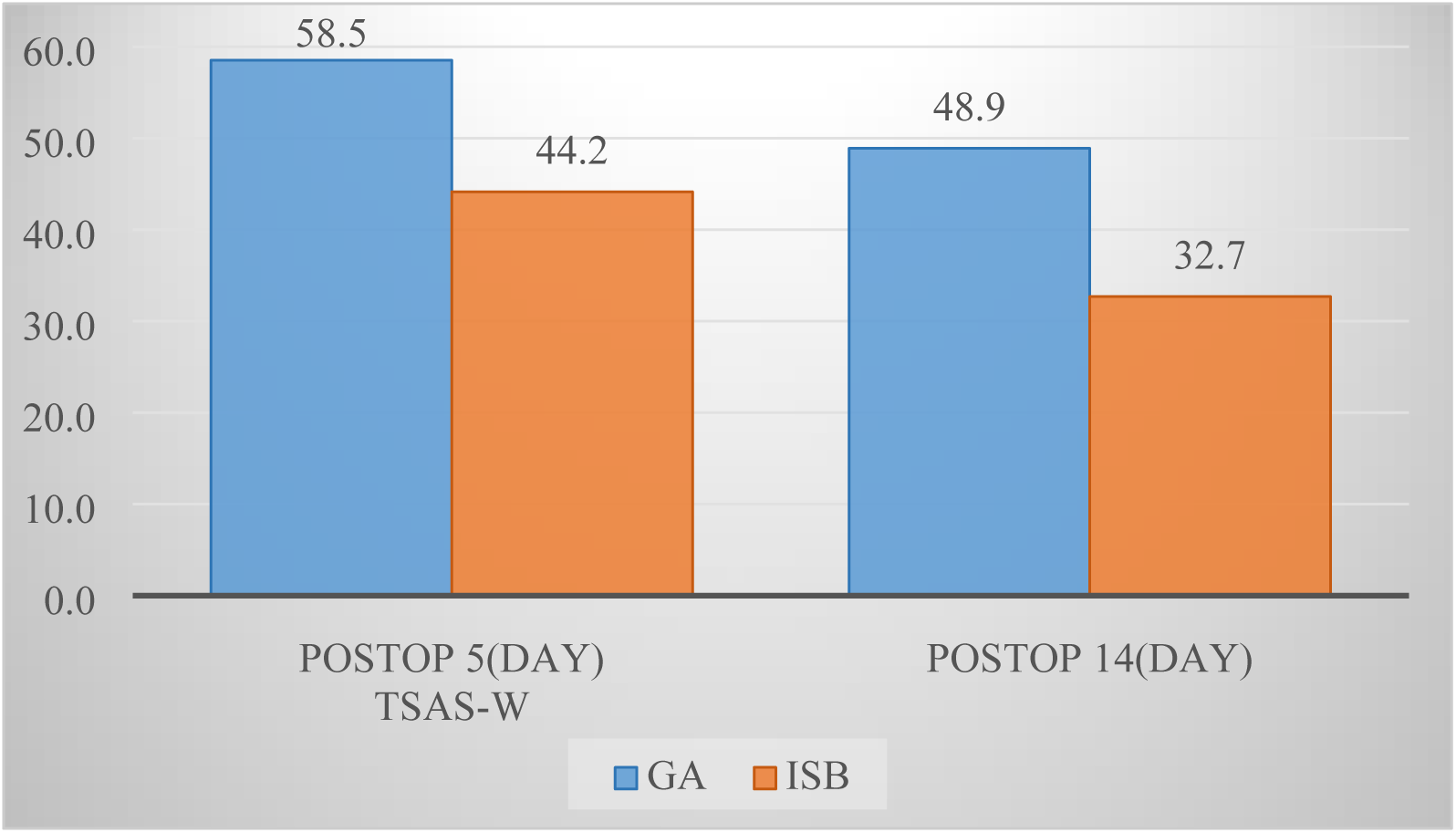
TSAS-W (Toronto Symptom Assessment System for Wounds) evaluation between groups.

**Table 2:**
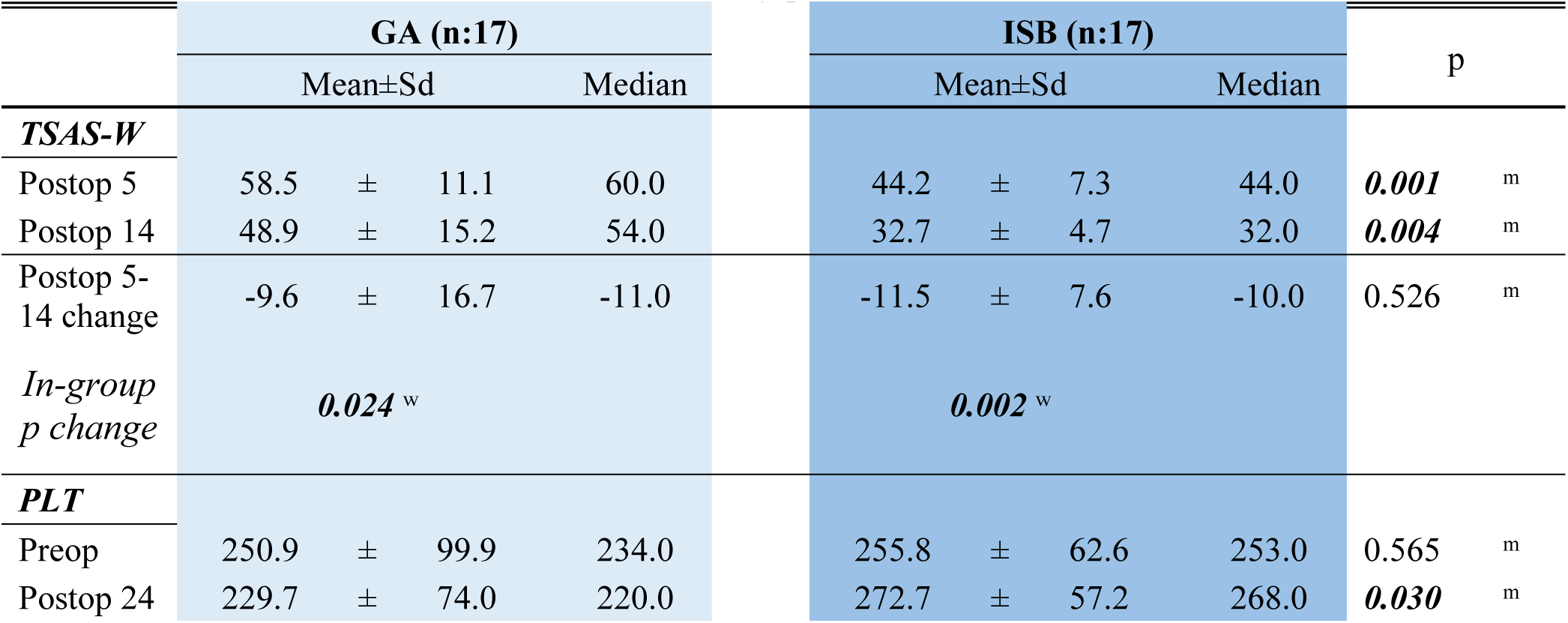
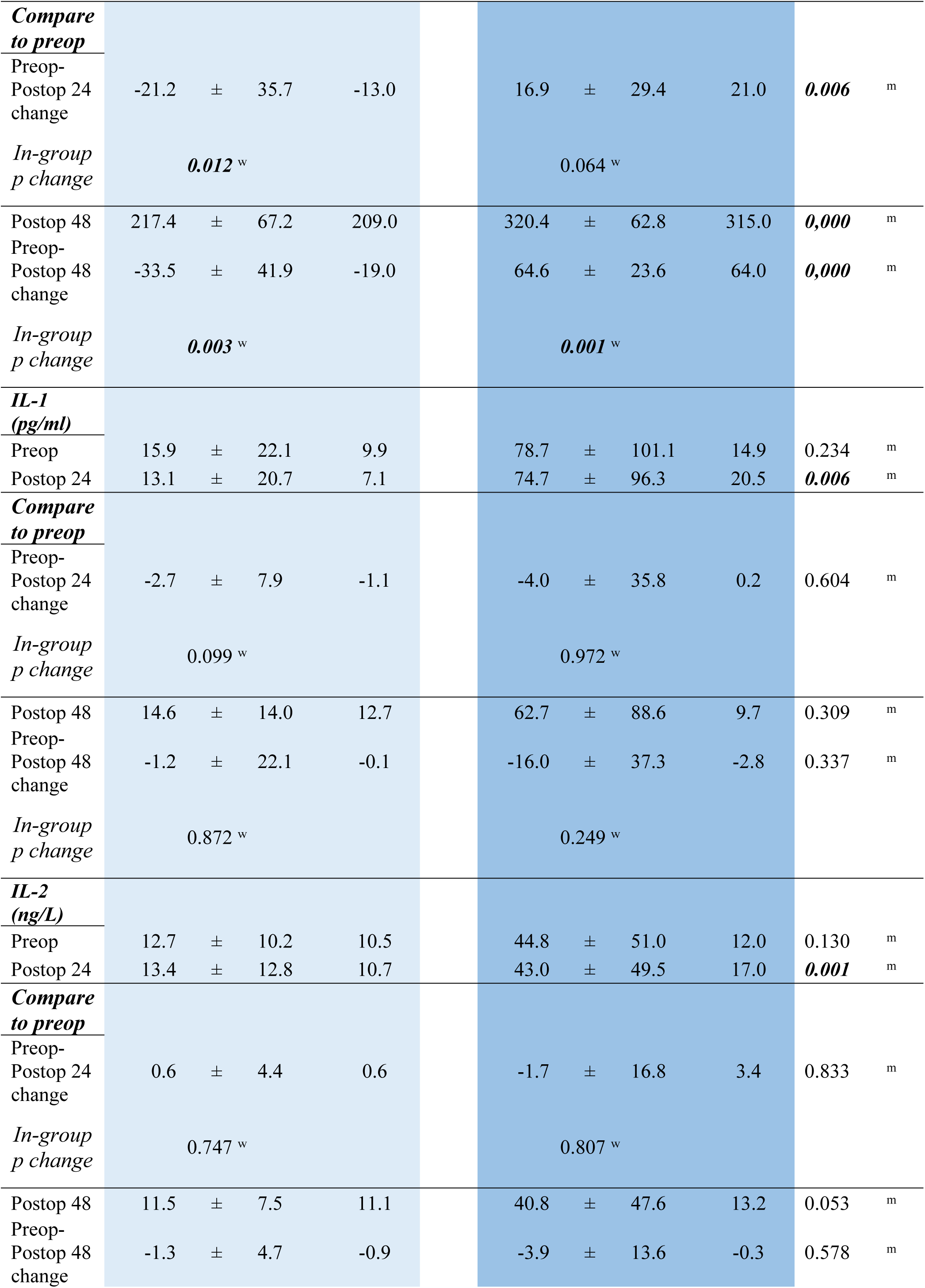
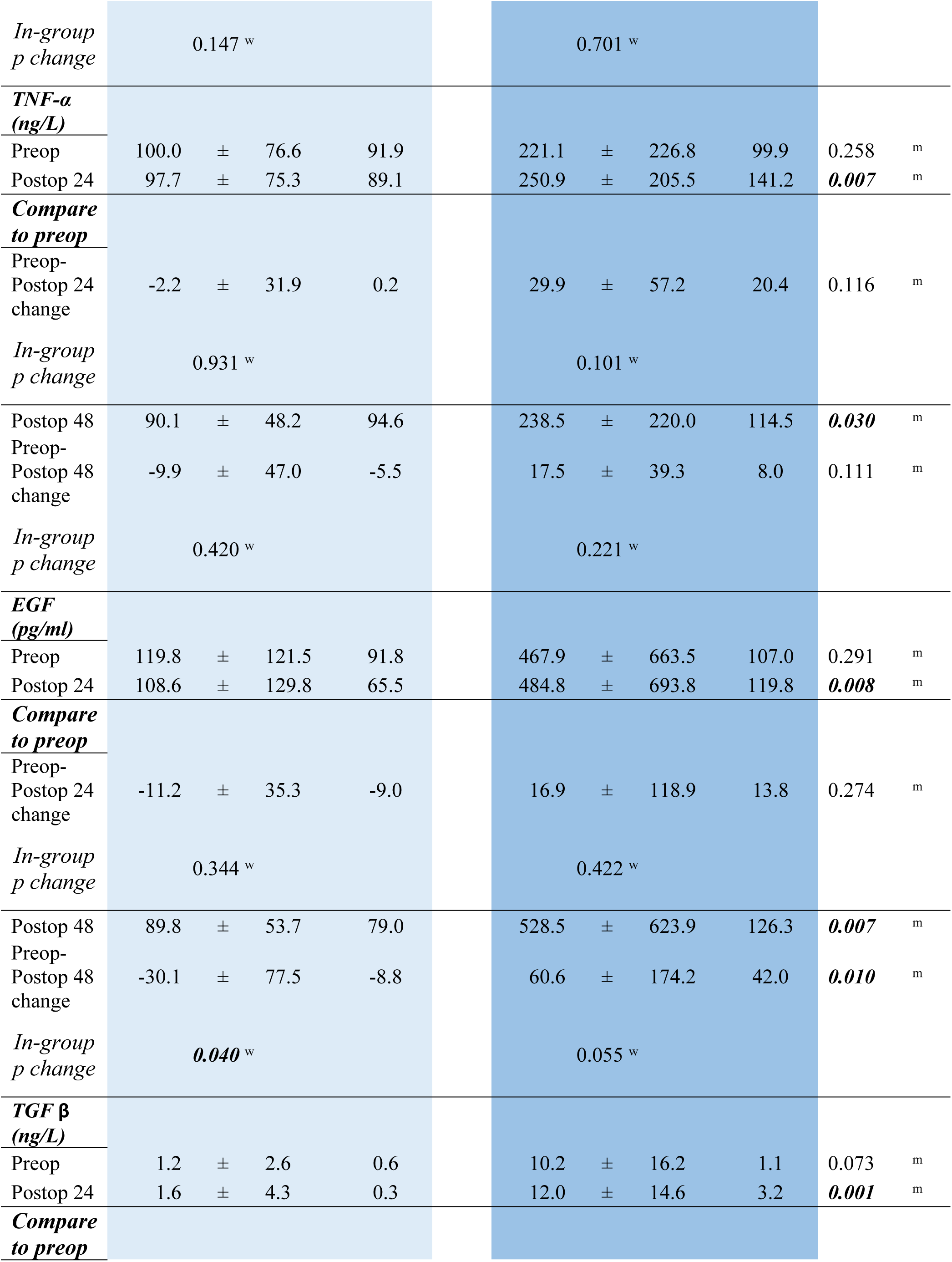
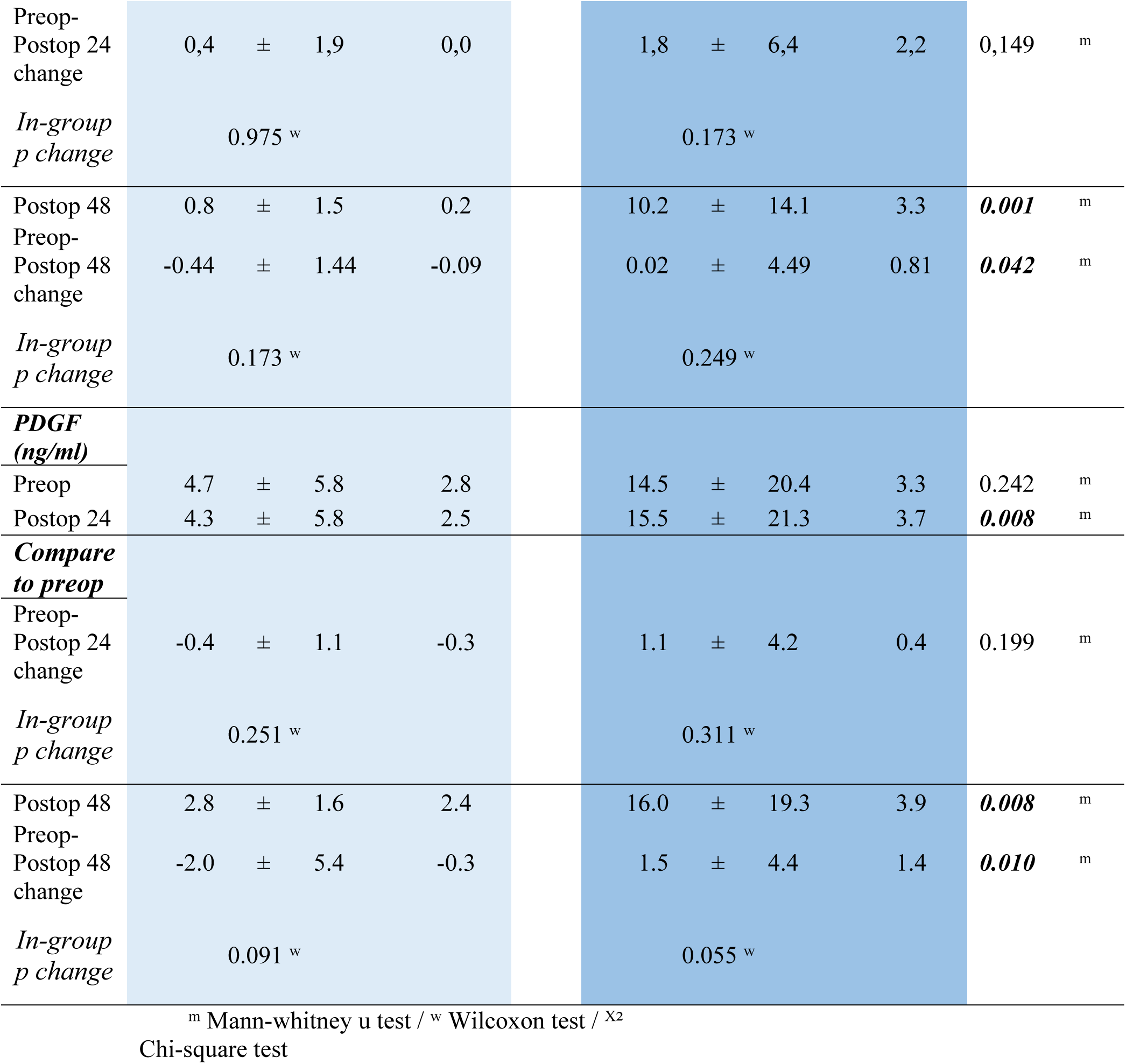
Changes in wound healing and immunity parameters.

When the groups were compared in terms of platelet levels, no significant difference was observed between the general anesthesia and interscalene groups in the preoperative values. However, postoperative 24th and 48th-hour platelet levels were found to be significantly higher in the interscalene group (p < 0.05). In the general anesthesia group, platelet levels did not differ significantly between the preoperative and postoperative time points. Although the interscalene group showed an increase in platelet levels at the 24th postoperative hour compared to preoperative values, this difference was not statistically significant. However, the 48th-hour postoperative platelet levels were significantly higher than the preoperative levels in the interscalene group (Table 2).

In the intergroup comparison of IL-1 and IL-2 levels, no significant differences were observed between the groups in preoperative and postoperative 48th-hour values (p > 0.05). However, at the 24th postoperative hour, both IL-1 and IL-2 levels were significantly higher in the interscalene group compared to the general anesthesia group (p < 0.05) (Table 2).

No significant intergroup differences were observed in TNF-α levels when preoperative and postoperative values were compared in both the general anesthesia and interscalene groups (p > 0.05). Similarly, intragroup analyses revealed no statistically significant changes in TNF-α levels between the preoperative and postoperative time points within either group (p > 0.05) (Table 2).

Preoperative EGF levels did not differ significantly between the groups (p > 0.05); however, postoperative EGF levels at 24 and 48 hours were significantly higher in the interscalene group compared to the general anesthesia group (p < 0.05). In intragroup comparisons, no significant differences were observed between preoperative and postoperative EGF levels in the general anesthesia group (p > 0.05), whereas the interscalene group showed significantly higher EGF levels at both 24 and 48 hours postoperatively compared to baseline (p < 0.05) (Table 2).

There was no significant difference in preoperative TGF-β levels between the general anesthesia and interscalene groups (p > 0.05) (Figure 3). However, postoperative TGF-β levels at 24 and 48 hours were significantly higher in the interscalene group compared to the general anesthesia group (p < 0.05) (Table 2).

**Figure 3:**
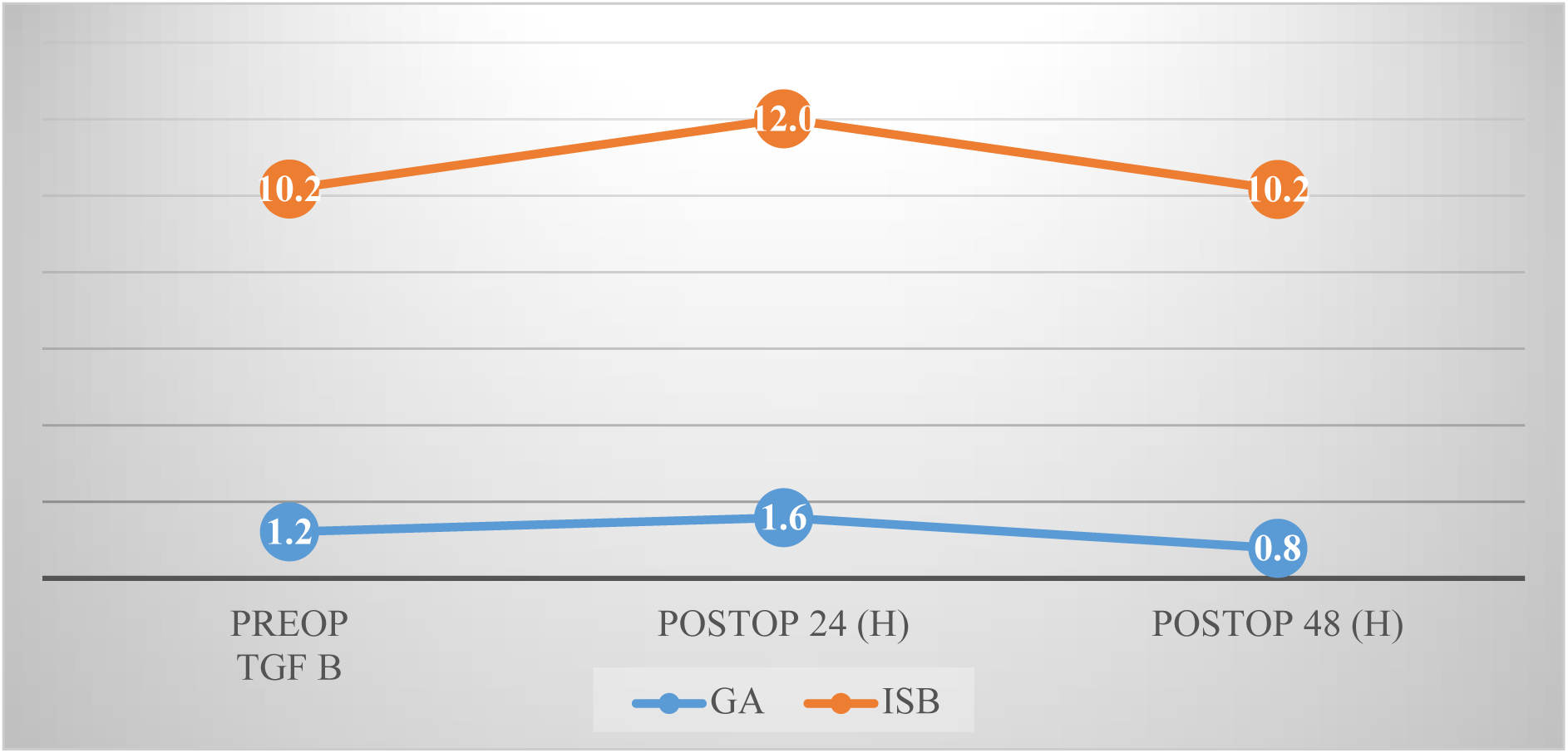
TGF β level changes.

There was no statistically significant difference in preoperative PDGF levels between the general anesthesia and interscalene groups (p > 0.05). However, postoperative PDGF levels at 24 and 48 hours were significantly higher in the interscalene group compared to the general anesthesia group (p < 0.05). In within-group analyses, postoperative 24- and 48-hour PDGF levels did not differ significantly from preoperative values in either group (p > 0.05) (Table 2). In the interscalene group, the VAS scores at postoperative 0, 4, 8, 12, 16, 20, and 24 hours were significantly lower compared to the general anesthesia group (p < 0.05). VAS scores at postoperative 4 and 8 hours did not differ significantly from preoperative values for interscalene group (p > 0.05). However, at postoperative 12, 16, 20, and 24 hours, the VAS scores were significantly higher compared to preoperative values (p < 0.05) (Table 2).

In the interscalene group, the mobilization times, the amount of postoperative analgesics used, and the incidence of postoperative nausea and vomiting were found to be significantly lower compared to the general anesthesia group (p < 0.05)(Table 3)

**Table 3:**
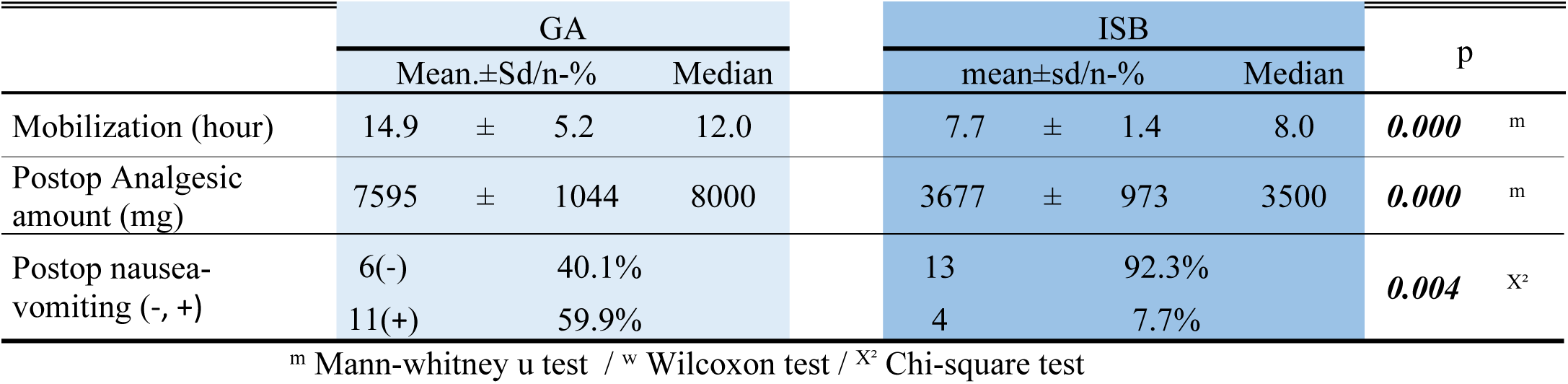
Postop parameters.

## Discussion

This study evaluates the effects of general anesthesia and interscalene block applications on wound healing, pain management, analgesic consumption, and postoperative complications. The study revealed significant differences between the general anesthesia (GA) and interscalene block (ISB) groups in terms of postoperative inflammatory response, analgesic efficacy, and wound healing. The finding that interscalene block application is more effective than general anesthesia and accelerates the recovery process in the postoperative period is consistent with the current literature. Interscalene block provided advantages, particularly in pain management, and reduced mobilization times and postoperative analgesic requirements. These results indicate that interscalene block is an effective analgesic method.

The study revealed significant differences in biomarkers such as platelet count, IL-1, IL-2, TNF-α, EGF, TGF-β, and PDGF between the interscalene and general anesthesia groups. The impact of these parameters on wound healing provides valuable insights into how different anesthetic techniques influence biological processes. It is hypothesized that the interscalene block positively impacts these biomarkers, thereby accelerating wound healing and improving the recovery process.

No statistically significant differences were found between the groups regarding demographic data, ASA scores, or smoking status, ensuring the validity of the comparison. Platelet counts, as indirect markers of systemic inflammation and tissue injury, significantly increased at 24 and 48 hours postoperatively in the ISB group. This may suggest a prolonged inflammatory response modulated by local immune mechanisms triggered by the block.

Interleukin-1 (IL-1) and Interleukin-2 (IL-2) levels showed no preoperative or postoperative (48h) differences between groups. However, both were significantly elevated at 24 hours in the ISB group, potentially reflecting an early immune activation caused by regional anesthesia and its influence on the local microenvironment (18, 19, 12).

Tumor Necrosis Factor-alpha (TNF-α), a central cytokine in inflammation, did not show significant differences between or within groups. This may indicate a limited role of TNF-α in the anesthesia-related inflammatory modulation in this surgical setting (13, 20).

Epidermal Growth Factor (EGF), a key mediator of epithelial repair, was significantly higher in the ISB group at 24 and 48 hours postoperatively, despite similar preoperative values. This suggests that ISB may facilitate early regenerative responses by reducing systemic stress and enhancing local perfusion (21).

Transforming Growth Factor-beta (TGF-β) levels were also significantly higher at 24 and 48 hours postoperatively in the ISB group. This cytokine is involved in both inflammation and tissue remodeling, and its upregulation may contribute to improved healing outcomes (22).

Platelet-Derived Growth Factor (PDGF) levels were significantly elevated in the ISB group at postoperative 24 and 48 hours, although no significant intragroup differences were noted. Given PDGF’s established role in tissue regeneration and fibroblast activation, its increase may support the superior wound healing observed in the ISB group (23).

VAS pain scores were consistently lower in the ISB group across multiple time points postoperatively. Especially during the early postoperative period (0–8h), VAS scores remained close to baseline in the ISB group, suggesting effective analgesia while the block remained active. These findings align with previous reports demonstrating ISB’s superior efficacy in early postoperative pain control (24, 25, 5).

Additionally, the ISB group experienced shorter mobilization times, reduced analgesic consumption, and lower rates of postoperative nausea and vomiting. These advantages have been widely reported and highlight the clinical value of regional anesthesia in enhancing postoperative recovery (26, 27).

Wound healing, evaluated using the TSAS-W scoring system, showed significantly better outcomes in the ISB group on postoperative day 5 and 14. Although both groups improved by day 14, the ISB group maintained significantly lower scores, reflecting enhanced healing. The observed elevations in growth factors and cytokines may contribute to these favorable results (28, 17).

In conclusion, interscalene block provides not only superior analgesia but also modulates systemic inflammation and supports improved wound healing compared to general anesthesia. These findings support the preferential use of ISB in appropriate surgical candidates, especially when early recovery and wound integrity are critical.

### Limitations

This study has several limitations that should be acknowledged. The relatively small sample size (n = 34) represents the primary limitation. However, this number was determined through an a priori power analysis, which demonstrated sufficient statistical power to detect large intergroup differences. The restricted sample size was also influenced by the relatively low incidence of elective open shoulder surgery and the intensive nature of serial biomarker assessments, which required strict standardization and repeated blood sampling. Furthermore, a narrow inclusion criterion was deliberately applied to obtain a highly homogeneous patient population, thereby minimizing potential confounding factors and increasing internal validity. Although the limited sample size may reduce the generalizability of the findings, the consistency of results across clinical and immunological parameters supports the robustness of the conclusions. Secondly, the anesthetic agent used was solely bupivacaine. It is important to consider that the effects of different local anesthetics may vary, and these differences could influence the outcomes. Additionally, the relatively small sample size (17 patients per group) represents a key limitation of this study. Furthermore, cytokine parameters were only assessed during the first 48 hours postoperatively, which limits the ability to evaluate long-term effects. The long-term impact of cytokines on wound healing and inflammation could have been better assessed by measuring these parameters at a later time point, such as on day 14. Another limitation is that, the study was conducted in a single center and involved a specific patient cohort. Future research could increase the generalizability of the findings by including patients from different centers and with various disease conditions. Moreover, since only bupivacaine was used in this study, future studies incorporating different local anesthetics and adjuvants could provide a broader dataset for comparison.

## Data Availability

Data cannot be shared publicly because of Patient confidentiality. Data are available from the Van Yuzuncu Yil University Institutional Data Access / Ethics Committee (contact via Şevin Kartal- Van Yuzuncu Yil University Ethical commitee assistant)) for researchers who meet the criteria for access to confidential data. The data underlying the results presented in the study are available from (Van Yuzuncu Yil University Department of Anesthesiology records).

https://www.yyu.edu.tr/Birimler/330

